# Malaria prevention in the age of climate change: A community survey in rural Senegal

**DOI:** 10.1101/2024.10.26.24316180

**Authors:** Andrew C.L. Sherman, Jesse D. Matthews, C. Andrew Aligne

## Abstract

**Background:** Malaria results in over 600,000 deaths per year, with 95 percent of all cases occurring in sub-Saharan Africa. Despite significant steady reductions from 2000 to 2015, there has been a recent resurgence. The estimated 2025 Africa death rate was recalculated to be 51.8 people per 100,000, whereas the previous estimate was 15.9. A potential explanation for this very significant setback is that increasing temperatures associated with global warming have made it more difficult to use insecticide treated mosquito nets. This study evaluated a rural west African population to determine barriers to mosquito net use, including heat and outdoor sleeping.

**Methods:** This study used a social ecological framework used by the Peace Corps to determine this community’s barriers to malaria prevention. We practiced community-based participatory research by developing and implementing a survey in rural southeast Senegal. Local village health workers were trained specifically to implement this survey. Observations of the mosquito nets and sleeping spaces were performed by surveyors. 164 households in 20 villages were surveyed from October to November of 2012.

**Results:** 164 of 164 selected households were surveyed, giving a 100% response rate, representing 21% of this local population. For the 1806 family members, respondents assessed a total need of 1565 nets, implying that each individual in this area needs 0.86 nets (95% CI: 0.77-0.95). The main reasons for not using an available net were heat and fragility of the nets. This population had very positive attitudes regarding mosquito nets and appreciated the work of local malaria educators.

**Conclusions:** The estimated need of 0.86 nets per person is 54% higher than the World Health Organization recommendation. Heat was found to be a major barrier in using a mosquito net, causing more people to sleep on outdoor structures. This study’s findings suggest the ratio of nets to people may need to be adjusted for the substantial increase in outdoor sleeping. Head of household responses in this population were found to have no systemic bias and would be an accurate way to assess a family’s need of nets. Deploying more malaria educators to this area would be appreciated and beneficial.

## Introduction

Malaria results in over 600,000 deaths per year globally. Approximately 95 percent of all malaria cases and deaths occur in sub-Saharan Africa, with 80% of those deaths in children under 5 years of age. Despite significant steady reductions from 2000 to 2015, there has been a recent resurgence. The estimated 2025 Africa death rate is now 51.8 people per 100,000, whereas the previous estimate for 2025 was 15.9 [1].

One potential explanation for this catastrophic setback is that increasing temperatures associated with global warming have made it more difficult to use insecticide treated mosquito nets (ITN), a proven strategy in the fight against malaria. Based on our experiences in villages in Senegal, we hypothesized that we would find that many people in rural areas spend considerable time sleeping outdoors, where it is somewhat cooler than inside their huts. Standard ITNs are designed for indoor use, so people sleeping outside generally lack protection from mosquito bites.

The goal of the study was to identify social and environmental factors that could be impeding malaria control by diminishing effective net usage. In addition to outdoor sleeping, we considered the knowledge and perceptions of malaria and ITNs, and community health priorities.

## Materials and methods

This study used a framework (Fig 1) used by the Peace Corps specifically to look at malaria control in rural sub-Saharan Africa [2]. This framework combines important aspects of previous models and incorporates a state of sustainability [3]. The Peace Corps framework has potential to guide effective interventions, based on a community’s assessment of barriers to regular ITN use.

**Fig 1.**
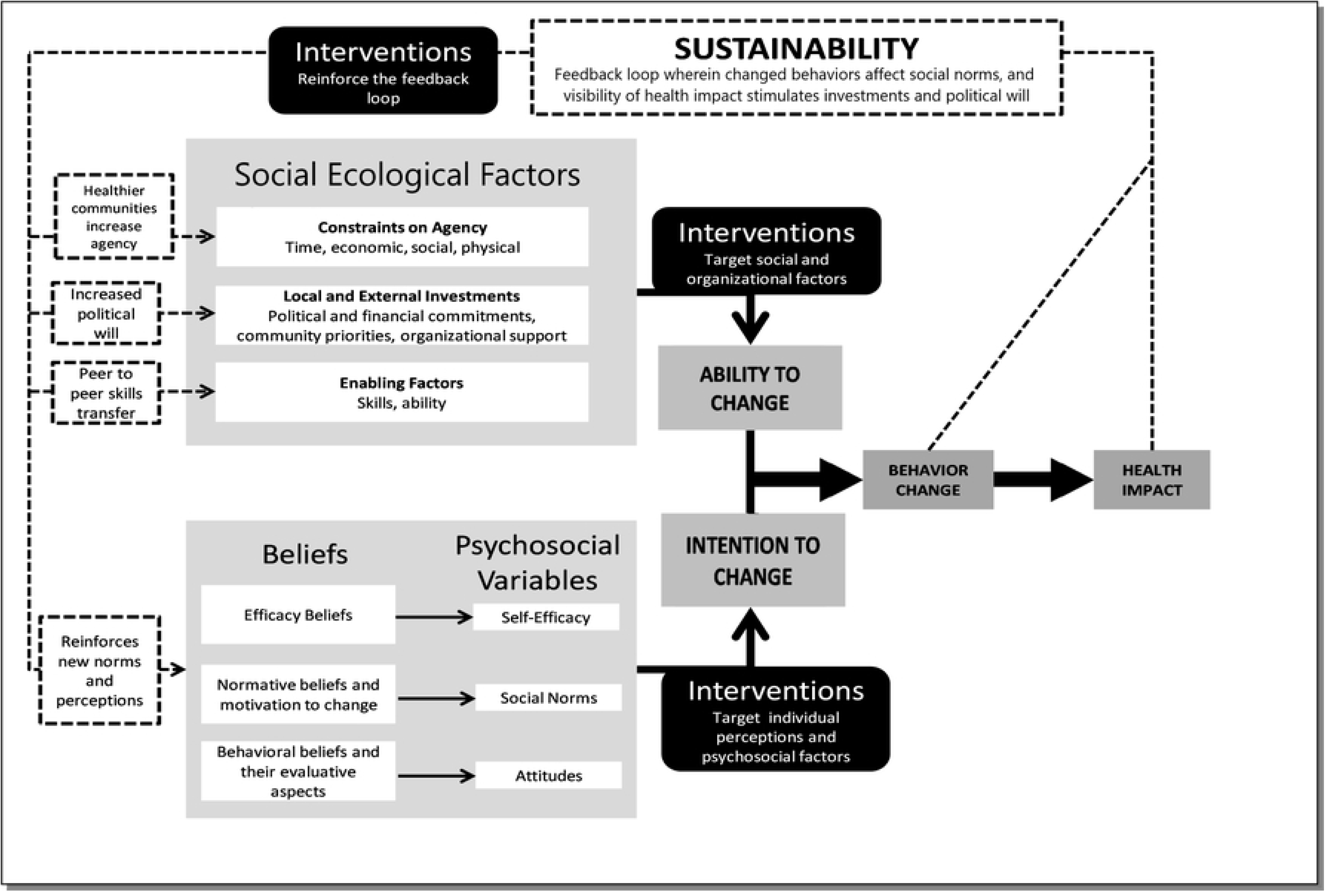
Study Theoretical Model. Adapted from McLaughlin and Peace Corps. This model integrates important social ecological factors and applies them to rural Senegal malaria prevention.

We practiced community-based participatory research by incorporating local stakeholders into the research team. Input from these community members informed the study’s design, implementation, analysis and interpretation. Our survey questions were adapted from the 2008-2009 Senegal Malaria Indicator Survey [4]. A range of open-ended questions were added to the survey to assess key components of behavior, with special attention to barriers of ITN use. The study questions reported here asked about the number of nets needed for everyone in a household to sleep under one, the motivations for people to use their nets, the reasons people may not use nets, ways the nets could be modified to improve use, other ways people believe malaria could be prevented, and possible opportunities for community level interventions to promote the use of ITNs. We also encouraged the surveyors to include observations and relevant comments on the survey. Survey questions are available for review in the appendix (S1-S3 Files).

### Study population

This study was conducted in the rural region of Kedougou (12.32° N, 12.18° W), located in the southeast corner of Senegal. The population of Senegal is 18 million, and the population of Kedougou is 245,000 [5,6]. The rainy season extends from May to November [7]. Kedougou has a poverty rate of 71.3% and an adult illiteracy rate of 58% [6]. Kedougou is composed of four sub-regions referred to as arrondissements. Our research focused on the arrondissement of Bandafassi and the 37 villages served by its health post (Fig 2). The village populations ranged from 30 to 952 people. This is a rural population, and most families have a livelihood of agriculture and cattle herding[5]. Over 95% of the population is Muslim, with the remaining practicing Christianity or animism [8]. The main ethnic groups in this sub-region are Peul, Mandinka, Bedik, and Bassari. French is taught throughout the school systems, so many residents communicate well in French. The average household size is larger than other areas in West Africa [9]. The standard household has 8 people and larger “collective” families average more than 15 people [5,6]. This area was selected because it has been a zone of high malaria morbidity and mortality.

**Fig 2.**
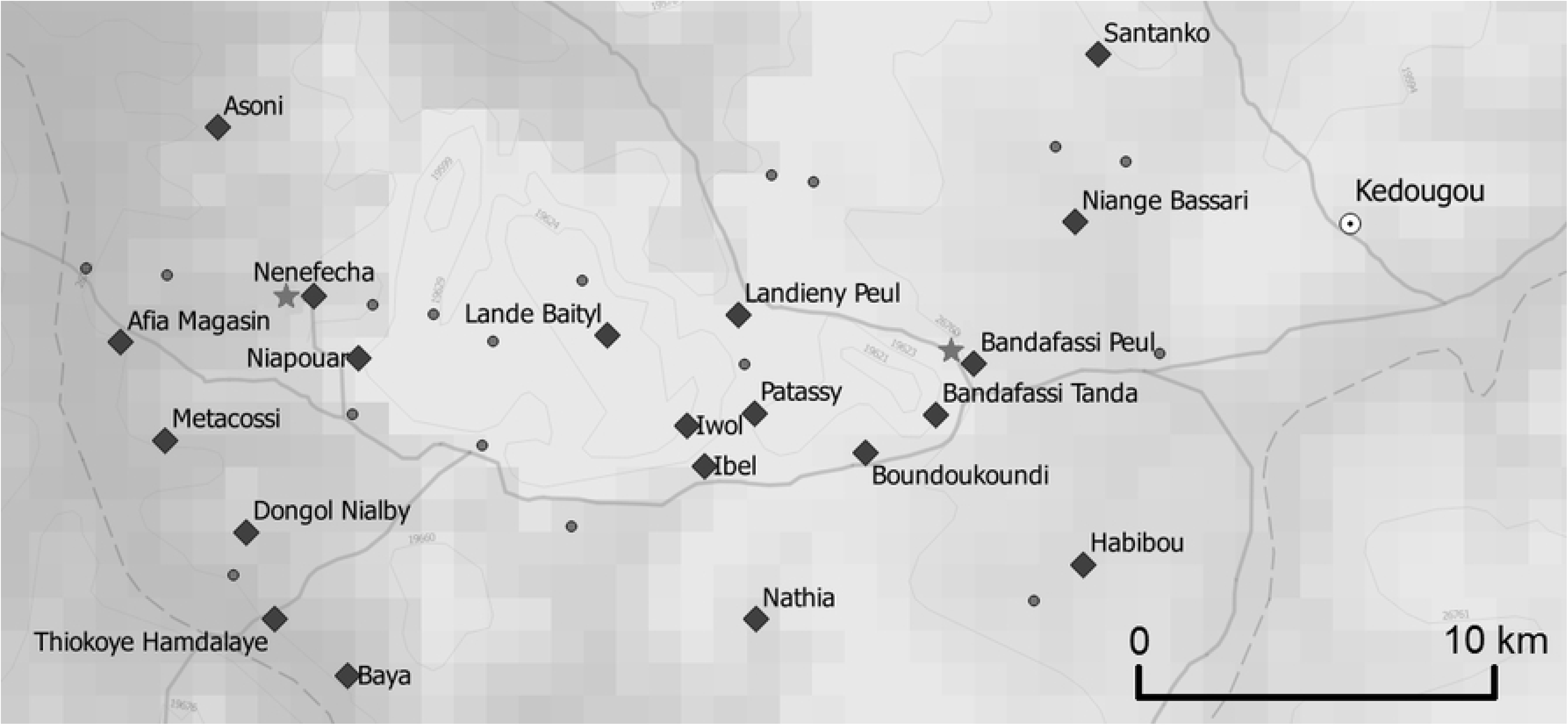
Map of the 37 Villages served by the Bandafassi health post in southeast Senegal. Black diamonds were selected villages. Small circles were unselected villages. Stars indicate a source of staffed health care access. (QGIS 2.4.0)

### Sampling and data collection

The study acquired preliminary census data from the Bandafassi health post to determine village population size. To confirm current population numbers, we also obtained village census data from each of the village chiefs served by the Bandafassi health post. We performed a simple random sample to select households, using a random number generator (Random~Number, PocketLab). We selected between 4 and 20 households per village, or between 15 to 33% of all households, depending on the size and location of the village. In all, we selected 164 households from 20 villages. Each questionnaire was labeled with a random unique identifier number.

In Senegal, the research team met with the coordinating Bandafassi health post nurse, as well as the village chiefs from sampled villages. The village chiefs agreed to partner with the study team, providing up-to-date census data and verification that questionnaires were properly administered by the surveyors.

The questionnaire was piloted in a village that was not part of the study sample. Based on feedback from piloting the survey, we adapted the questionnaire for local comprehension and cultural relevance. The study used local surveyors, recommended by the health post nurse, to administer the survey. Surveyors were selected based on their experience with similar surveys and their bilingualism in French and a local language. The study employed eleven surveyors – 7 men and 4 women. The research team and health post nurse developed a training curriculum for the surveyors. After the training, the surveyors demonstrated enhanced abilities to implement the questionnaire in a culturally sensitive manner.

The surveyors conducted 164 structured interviews between October 31^st^ and November 30^th^ of 2012. Data collection coincided with the last months of the rainy season, when at-risk people could have been decreasing their net use. Surveyors were assigned to villages according to their language skills. Surveys were conducted in the homes of participants and directed at the heads of households, with input from available family members. Participants were not paid.

### Data analysis

Surveyors recorded responses for each questionnaire verbatim in the language used by the participant. If a response was in a local language, the multilingual interviewer would immediately translate and write the response again in French. Translations were made from French to English by a bilingual research team member. English translations were transcribed into a REDCap interface. For descriptive analysis of responses to open-ended questions, three researchers reviewed all responses and came to a consensus on which categorical codes best represented the response themes. The codes were also chosen based on their potential to reflect known categories of ITN-use barriers and the themes of the selected theoretical framework (Fig 1). Analysis was used to address the gaps in the present literature, specifically the knowledge, perceptions and cues to action of ITN use. Four independent researchers assigned codes for each survey response. The team discussed disagreements until a consensus interpretation was agreed upon.

A section designed to quantify family members and sleeping spaces from the head of the household and surveyor observations was also evaluated. The research team made field observations of mosquito nets and village beds. Surveyors were instructed to estimate that one ITN was needed for each sleeping space. Members took detailed notes on these observations, and then repeatedly met to discuss possible interpretations. Senegalese members of the research team verified interpretations within the context of the local culture. Incomplete responses were excluded from the analysis.

### Ethical considerations

The University of Rochester Research Subjects Review Board approved this project. The Bandafassi health post administration approved and partnered with this project for the training of the surveyors. We met with all village chiefs of the selected villages, and they approved of this project. Each participant gave verbal consent before the questionnaire. No personal identifiers were recorded.

## Results

164 of the 164 selected households were successfully surveyed, giving a 100% response. Most individual questions had responses over 96%. The question with the lowest response had a 93% response. The 164 households represented 1806 people or about 21% of the total population of the Bandafassi region. 152 heads of household gave updated information on household size.

The 20 selected villages were an average of 9.7 kilometers away from the nearest staffed health care access. The average distance from health care access for all 37 area villages was 9.3 kilometers. The 20 selected villages had an average population of 350 people. The average population of all 37 area villages was 235 people. The average household size was 11.9 people.

### Perceived need for nets

97.6% of households had access to at least one net. For the 1806 family members, respondents assessed a total need of 1565 nets. This implies that each individual needs 0.86 nets (95% CI: 0.77 – 0.95), i.e. the 12,441 people in this region need a total of 10,678 nets.

### Barriers to optimal net utilization and malaria control

#### Heat

When asked “When you think about mosquito nets, what do you dislike about them?” heat or lack of airflow was commonly discussed.

> *‘When the weather is hot, it keeps the air from getting through and makes it hard to sleep comfortably.’*

> *‘…sometimes one has the impression of sleeping in a hole.’*

> *‘[T]he mosquito nets make it hot and suffocate us.’*

When asked “There are people that do not use a mosquito net every day. In your opinion, why do these people not use a net every day?” a respondent discussed the mosquito net’s effect on airflow:

> *‘During the hot weather, some people say that the mosquito net makes it even hotter.’*

When asked “How could the mosquito net be improved?” another respondent discussed how outdoor sleeping can cause accelerated damage of nets:

> *‘If it’s in the room set up properly under the mattress then it will be fine. But if it’s outside with the beds that don’t have mattresses, then it will deteriorate quickly.’*

When asked if they had anything to add, this respondent discussed a sense of vulnerability with being outside in the evening:

> *‘Mosquito nets are effective. Except that when we are outside before going to bed, the mosquitos that bite us give us malaria.’*

A surveyor observed situations where children were vulnerable during outdoor sleeping:

> *‘There were miradors* [elevated sleeping platforms] *outside but no mosquito nets. Some children were sleeping there. Others on the other hand were sleeping inside where all the beds had nets hanging.’*

#### ITN fragility

A majority of respondents described ITN fragility as a reason for needing more nets. When asked: “How could the mosquito net be improved?” 62.8% of respondents discussed a need for increased durability:

> ‘*They need to be made tougher and longer-lasting.’*

> *‘We have cots made of pleated wooden slats; we would like the nets to be improved so that they are more appropriate for our beds.’*

Also, when asked: “Are all the mosquito nets you received at the last distribution still here?” 23.7% said no. When a follow up question asked: “What happened to the other nets?” 69.1% mentioned damage.

Some respondents suggested improvements with ITN design. Many of these ideas involved reducing ITN damage by customizing them to the needs and preferences of the population:

> ‘*We would like mosquito nets that have doors so that if someone or something comes into my pasture to attack my animals that I might be able to leave easily to go to their rescue.’*

#### Overwashing

When asked what happened to the missing nets, respondents described concerns about how the washing process could damage the nets:

> ‘*The mosquito nets lasted, but then got ruined. The women wash them and dry them under the hot sun and so they wind up getting ripped.’*

Regarding the drying process, nets “should be stored properly in dry season”, and after washing:

> *‘…they shouldn’t be hung up on wood.’* When asked “How could the mosquito net be improved?” 22% of respondents mentioned washing the net. Respondents regularly emphasized the importance of cleanliness, which was often linked to increased rates of net washing:

> *‘If the mosquito net is dirty, we wash it and hang it back up.’*

Some respondents viewed the chemicals as something that causes illness and thus needed to be cleaned or removed:

> *‘There are mosquito nets that have an odor that causes colds, so we wash them before using them.’*

#### Isolation from health services

Respondents reflected on how they are far from health care services. The distance to a health care provider was commonly mentioned. At the end of the survey, a respondent was asked if he wanted to add any other comments:

> *‘To travel to the other villages when somebody is gravely ill, you can imagine is very risky.’*

Surveyors from this region regularly discussed the burdens of disease and the lack of health care access:

> ‘*These people are really exhausted from going from village to village to get access to care or even just to get medical information.’*

> ‘*We need someone to come and help the people who live far from the city because they face enormous barriers to receiving care and they need to be educated on how not to get sick.’*

### Facilitators of net usage and malaria control

#### Acceptability of community malaria educators

When discussing other community priorities, participants often valued the role of education. When asked “If there where health aides who were trained to help solve problems with mosquito nets and malaria in general would you be interested in having someone come to the household to help you?” 98.2% said yes. 91.3% thought this malaria educator should visit their homes two or more times a year. 52.8% thought they should visit 4 or more times a year, regularly suggesting even more frequent visits from “*three times during the rainy season*” to “*all the time*”.

Here, a respondent shows an appreciation for this study’s survey as an anti-malaria intervention, also demonstrating how past education has improved the local knowledge base:

> *‘I am very happy about this visit that is very important for us. It is a great pleasure to see you in my house to do something about this mosquito problem … Mosquito nets protect us from mosquitoes, which give us malaria. Malaria is a very serious disease transmitted by the female of a mosquito called anopheles.’*

When asked “In your opinion, what do you think the people of the village could do to prevent more cases of malaria? You can talk about things other than mosquito nets”, education was discussed regularly:

> *‘What the people of the village could do to help prevent malaria is to organize educational sessions and conversations all the time.’*

> *‘The people of the village need to organize … discussions on malaria and ways to prevent it.’*

> *‘We should have conversations every month, if that’s possible. Especially before, during, and after the rainy season.’*

#### Acceptability of ITNs

When asked “When you think about mosquito nets, what do you like about them?” 100% of respondents expressed positive attitudes toward nets, including protective benefits and comfort. They also gave examples of perceived risks of malaria:

> ‘[The net*] protects our health a lot. If you sleep under a mosquito net, you will not get bitten by a mosquito or other insects. It decreases dust. It is very important because it protects us against lots of diseases.’*

> *‘[H]anging up mosquito nets is better than having to throw away all of one’s money on prescriptions.’*

> *‘If we didn’t have mosquito nets, we would have people falling ill all the time; that is why I always tell the children to lower the mosquito nets when they go to bed.’*

When asked “There are people that do not use a mosquito net every day. In your opinion, why do these people not use a net every day?” 63.1% mentioned the words “negligence or ignorance”:

> *‘People who don’t use a mosquito net every day in my opinion are people who don’t know the risks they are taking.’*

> *‘People who do not use mosquito nets every day, in my opinion, it is because of laziness or negligence…’*

Respondents also had trouble imagining or explaining why people who live in their culture would not use a net:

> ‘*In my opinion, it is the Westerners who do not use mosquito nets. In Africa, there’s no way to live without insecticide.’*

> ‘*I never heard anyone here say that they dislike mosquito nets.’*

#### Suggestions from the community

When asked about possible community actions to reduce malaria, interventions about cleaning were by far the most common response (60.4%).

> *‘The people of the village could help with the prevention of malaria by cleaning up the village properly.’*

> *‘The village residents must organize themselves to fight against filth…’*

> *‘Avoid pools of stagnant water, destroying them during the rainy season. Everything that could contain water or otherwise bring mosquitoes should be destroyed.*’

## Discussion

In a rural Senegalese population with a high malaria burden, our survey found a need of 0.86 nets per person. This is 54% higher than the current WHO recommendation of 0.56 nets per person [10]. The WHO recommendation for the number of nets per person has existed since 2010. The WHO suggests the “ratio can be adjusted as needed if there are data that support such adjustment”[11]. It has been suggested that the ratio calculation may need to be adjusted for the early damage of nets and its effect on ITN retention. This study’s findings suggests that the ratio should be adjusted further to account for the substantial and increasing degree of outdoor sleeping: to prevent malaria deaths, people need more nets than they are currently receiving. We believe this troubling finding is valid.

A major strength of this study is that the lead researcher lived in this area as a Peace Corps volunteer and established positive relationships with community leaders. These connections continued for many years via the public health work of the non-profit organization, Netlife. These past partnerships and the use of local languages helped create a strong, multifaceted study team. Survey piloting helped greatly to create a culturally accepted final version. The trust from these past relationships and the culturally sensitive administration of the surveys contributed to the 100% response.

The flipside of these relationships is that, as with any survey, there is the risk that participants will try to please the interviewer with their responses. However, we found many responses with negative impressions of nets, indicating a balanced impression. Another potential limitation is that our data for the number of nets needed by families was based on the head of household’s assessment. This is not as objective as counting all household members and sleeping spaces. There can be a concern that heads of household will ask for more nets than necessary. To address this, we analyzed a subset of open-ended responses from the surveyors. The surveyors had an option to write down observations of the sleeping areas. This wasn’t done by all the surveyors, but the responses that were documented helped to compare the number of requested nets versus the number of observed and verified sleeping spaces.

As the quotes below illustrate, observations by surveyors indicated that some villagers underestimated their need for nets, while others overestimated

In the first example, the surveyor’s observation suggests that there are 17 total sleeping spaces here, and the head of household stated a total need for only eight nets; this is therefore an underestimate.

> ‘[The head of household] has 9 inside beds with mosquito nets, of which 5 are hung up, 3 torn and 2 repaired. On the interior floor, there are 5 [sleeping spaces] that don’t have mosquito nets. Outside are 3 beds without mosquito nets.’

In contrast, the next example illustrates a head of household assessing a need for seven nets, whereas only three sleeping spaces were observed, thus suggesting an overestimate.

> ‘[The head of the household] *has a very well organized and very knowledgeable family since they answered all the questions easily and the house is very tidy. There are 3 interior beds all of which have mosquito nets hanging and the mosquito nets are not torn.’*

Overall, these responses showed some underestimates, some overestimates and many appropriate estimates of requested nets. So, we found there was no systematic bias of heads of household overestimating their families’ ITN needs.

Head of household assessment is a valuable tool in this population when it is hard to account for all household members and indoor/outdoor sleeping behaviors by briefly inspecting a home in the daytime. A head of household will know how members move from bed to bed within the family and its environment.

Another potential limitation is that our survey was conducted in 2012 and so could now be outdated. However, members of our team have visited these villages multiple times since then (as recently as 2023), and if anything, the situation with heat and outdoor sleeping has become more pronounced, and we believe it now requires urgent attention. We have made regular observations of outdoor sleeping. Most household compounds have outdoor structures used during the day for shade or for food preparation. During the evenings, many family members drink tea and talk on these structures. Because of the heat, many people prefer to sleep the first part of the night outdoors, because there is better airflow outdoors than in the huts. When cool enough, most family members wake up and move to the sleeping spaces inside the huts. Regularly, family members would sleep one-third to one-half of the night outside. Although we observed mosquito nets being used on some outdoor structures (S1-S4 Figs), this was rare. Most household members only have enough nets for their indoor sleeping spaces.

Because outdoor sleeping increases the number of sleeping spaces used by each family, it increases the number of nets needed to provide protection from mosquitos. In addition, if nets are used outside, they wear out faster. For example, the structures used for outdoor sleeping tend to have many sharp points, and nighttime winds cause the nets to regularly catch on these points. (S4 File) Very few ITNs are truly designed for outdoor spaces [12,13]. Consistent with Killeen and Grand Challenges in Global Health, this study supports the need to expand interventions to the outdoors [13,14].

Prior to this survey, few studies had specifically investigated outdoor sleeping as a barrier to ITN use. Studies conducted near Lake Victoria and in Burkina Faso found that outdoor sleeping was infrequent [15,16]. In contrast, we found outdoor sleeping to be extremely prevalent, consistent with reports from Niger and Iran [17,18]. Heat has been found to be a significant barrier to ITN use, and causes this study population to regularly sleep outside [19–22].

As in previous studies, this population demonstrates concerns about the durability of nets [23–27]. Study responses and our observations indicate that village bedframes often have jagged edges that increase the risk of rips in ITNs. When ITNs get dirty, people will wash the nets to improve their appearance in the home. However, some of the washing and drying techniques are abrasive and it is likely that many nets are overwashed. Previous studies verify that misunderstandings about ITN washing can damage the nets [28–32]. Future alterations of ITNs or community interventions that decrease over-washing will increase durability of the nets.

Although there is a large body of research that focuses on ITN uptake and ownership-to-use rates, there are few studies reporting on the development of ITNs as a social norm [3,19,33]. This study’s responses were saturated with positive comments about nets. Participants demonstrated knowledge about the deadly nature of malaria, the vulnerability to children, and the economic benefits of avoiding illness. When asked to come up with a reason people would not use a net every night, they often used the words “negligence” or “ignorance”. Research shows that when most members of a population shift to the disapproval of a previously common behavior, it represents a change in the social norm [34].

One factor in this level of success in community engagement to fight malaria is PECADOM PLUS (Prise en charge à domicile), which is a Senegalese Ministry of Health developed program that Peace Corps volunteers have extended into the rural communities [35]. PECADOM PLUS uses the village health leaders in an early detection system of new malaria cases. Many villages now have designated malaria champions who have participated in vaccine campaigns, ITN distributions and educational sessions. In the past, community participation like this has been found to reduce disease burden, if the champions are provided with appropriate resources, training and supervision [36–40]. Maintaining a high level of community involvement is also very important to sustain high levels of ITN use where malaria rates are decreasing [41–43].

## Conclusions

In a rural Senegalese population with a high malaria burden, our survey found a perceived need of 0.86 insecticide treated nets per person. This is 54% higher than the current WHO recommendation of 0.56 ITNs per person [10]. Moreover, our field observations indicated that the number of nets needed for adequate protection against malaria could be even greater.

One particular concern is global warming. Heat is a major barrier to the use of an available indoor mosquito net. Increases in temperature are likely to cause people to sleep on outdoor structures more often. If there is an increase in the already high prevalence of outdoor sleeping, then the number of needed nets will increase. We should provide more nets for people who sleep outside, and research should be conducted to develop modified nets more suitable for outdoor use. The reason people sleep outside is the heat, and this is likely to worsen with global warming. Therefore, it seems that efforts to decrease global CO_2_ emissions would help with malaria control.

Fortunately, this study also highlighted the presence of many assets for fighting malaria in this hyperendemic region. The population is already very well informed about malaria, engaged in ITN usage, and eager to keep learning about malaria control. An existing program of community health workers is well received. Thus, a feasible and acceptable intervention would be to deploy more local malaria champions who can provide regular educational updates on malaria and ITNs, identify and treat early cases of malaria, and empower communities by leading discussions on how local stakeholders can create and develop effective interventions. Increased malaria control services should especially be provided to families who live far from health posts or hospitals. Strengthening the village health worker network would be an inexpensive, highly effective method to reduce malaria deaths, as well as create an improved health care infrastructure to detect outbreaks of other diseases such as meningitis or Ebola.

This population places a high value on village cleanliness as a part of public health. It is therefore important to incorporate cleaning activities into community malaria interventions. With specific guidance, such programs could include activities like eliminating standing water, fixing doors or adding window screens, all of which would reduce the transmission of malaria. Hanging up ITNs as part of the home improvements and educating about over-washing would improve utility of the nets.

Implementation of the solutions suggested here should help accelerate the decline of malaria mortality in sub-Saharan Africa.

## Data Availability

All surveys (without any identifiers) are scanned and in a secure database. These can be made public, if needed.

## Acknowledgments

This work could not be done without the lifelong efforts of Bandafassi health post nurse, Mactar Mansaly. Thank you to all the assisting professionals at the University of Rochester Department of Pediatrics and the Hoekelman Center. Thank you to the Bandafassi area village health workers, the local Peace Corps Volunteers, the members of the Programme National de Lutte contre le Paludisme, and the Senegal Ministry of Health.

## Supporting information

**S1 Fig. Children at shade structure.** An outdoor shade structure, used during the day for food preparation. In the evening, this structure was a hub for making tea and conversation. For the first half of the night, this structure was regularly used for sleeping.

**S2 Fig. Outdoor sleeping space.** A rare example of outdoor structures with mosquito nets. These elevated sleeping platforms were made with long wood sticks, which typically had sharp ends which catch and tear ITNs.

**S3 Fig. Boy by outdoor sleeping space.** Another example of a typical outdoor structure made with bamboo-like grass. Here, the sharp ends of this material are evident.

**S4 Fig. Outdoor structure used for food preparation.** This outdoor structure (left of the hut) was commonly used for drying grain on its top level. Its top level was made from a piece of local wooden fencing, also with sharp edges.

**S1 File. Survey tool (French).** After piloting, the French version of this study’s survey was created.

**S2 File. Survey tool (English).** The English version of the survey, back translated from French.

**S3 File. Survey tool (Pulaar / Fulani).** The local language translation of the study survey.

**S4 File. Video outdoor sleeping.** Discussion of outdoor sleeping space usage

## Notes

### Competing Interest Statement

The authors have declared no competing interest.

### Funding Statement

The author(s) received no specific funding for this work.

### Author Declarations

This study was approved by the Research Subjects Review Board at the University of Rochester Medical Center, number RSRB00041566. Verbal consent was informed via a script utilized by Rollback Malaria Monitoring and Evaluation Reference Group, World Health Organization, United Nations Children's Fund, MEASURE DHS, MEASURE evaluation and the U.S. Centers for Disease Control and Prevention, 2005 Malaria Indicator Survey. The verbal script was also approved by the University of Rochester Medical Center's Research Subjects Review Board. The surveys had no identifiable information and unique identity numbers, ensuring confidentiality and making this survey minimal risk.

